# A protocol for assessment of interventions using a computational phenotype for Long COVID

**DOI:** 10.64898/2026.03.26.26347671

**Authors:** Amitabh Gunjan, Lawrence Huang, Anudeep Appe, Paul A. McKelvey, Heather A. Algren, Mark Berry, Essy Mozaffari, Bill J. Wright, Jennifer J. Hadlock, Jason D. Goldman

## Abstract

**Background:** Long COVID presents with one or multiple symptoms or diagnosable conditions after SARS-CoV-2 infection. To study whether use of the antiviral remdesivir in persons hospitalized with acute COVID-19 is associated with reduced Long COVID, we created a computational phenotype for Long COVID.

**Methods:** In electronic health records (EHR) from a multistate healthcare system (US), hospital admissions from 5/1/20 – 9/30/22 were reviewed. The study group was hospitalized with acute COVID-19 and the control group was hospitalized for other reasons without prior SARS-CoV-2 infection. The populations were balanced with overlap weights based on a high-dimensional propensity score of pre-specified variables and the top 100 comorbidities differing between the groups. Hazard ratios (HR) were calculated for the combined primary outcome: U09.9 (Post-Covid Conditions) or any incident secondary outcome from 90 to 365 days after admission. Secondary outcomes included 27 individual incident diagnoses, corrected for multiplicity with Holm-Bonferroni.

**Results:** Admissions included 45,540 with, and 409,186 without COVID-19 during the study period, evaluable for the primary outcome. After weighting, standardized difference was < 0.01 for all measured confounders including demographic and clinical features. In the COVID+ and non-COVID groups 38.0% and 29.3% met the combined primary outcome, respectively. Weighted HR (95%CI) for the primary outcome was 1.37 (1.35, 1.40), p < 0.0001. All secondary outcomes were associated with the COVID+ group, when adjusted for multiplicity. Incident diagnoses with strong associations (HR > 2) included thromboembolism, hair loss, diabetes mellitus, obesity, and hypoxia. Anosmia/dysgeusia was associated with COVID, but wide confidence intervals reflected few charted diagnoses.

**Conclusions:** Manifestations of Long COVID at population scale are detectable as part of routine symptoms and clinical diagnoses in the EHR after admissions for COVID-19, compared with all other hospital admissions. This *a prior* computational phenotype for Long COVID will be used to assess whether remdesivir use is associated with decreased Long COVID.

## Introduction

Long COVID is a major public health problem, estimated to affect approximately 15 million adults in the United States alone.^1^ Long COVID is difficult to define, as many of the clinical features are common symptoms and diagnoses in the general population and can occur for reasons other than as sequelae after SARS-CoV-2 infection. Recently, the National Academies of Science, Engineering and Medicine (NASEM) defined Long COVID as an infection-associated chronic condition that occurs after SARS-CoV-2 infection and is present for at least 3 months as a continuous, relapsing and remitting, or progressive disease state that affects one or more organ systems and can manifest in multiple ways including with symptoms or diagnosable conditions.^2,3^ The NASEM definition is intended to be a broadly applicable definition, and that subsets would need to be specified for research purposes. Here, we apply the concepts of the NASEM definition to measuring Long COVID outcomes in EHR data. We start with the approach used by Al-Aly^4^ assessing >800 high dimensional outcomes and refine via literature review of common features of Long COVID to a single combined primary outcome of Long COVID and a smaller set of individual secondary outcomes which are the components of the single combined primary outcome.

The pathophysiology of Long COVID is still undefined, but may relate to tissue injury, ongoing inflammation, autoimmunity, persistence of SARS-CoV-2 viral reservoirs, reactivation of other viruses or other contributions of the microbiome or virome, endocrine disruption, psychosocial factors, or a combination of these or other mechanisms.^5,6^ SARS-CoV-2 persistence may be a central hypothesis which could tie in other mechanisms, such as ongoing inflammation.^7,8^ Use of antivirals to disrupt SARS-CoV-2 persistence with either a prevention^9^ or treatment^10^ approach is being actively studied.^11^ Using the Long COVID outcomes developed herein and the analytic approach we develop in this protocol manuscript, we will study the effect of the antiviral remdesivir when given during acute COVID on the association with the subsequent development of Long COVID. This retrospective study is intended to provide non-causal inference as to whether remdesivir use during acute COVID-19 could be preventative for Long COVID.

This retrospective study is performed in two stages. In stage 1, we pre-specify the outcomes of Long COVID in a cohort of hospitalized persons with or without acute COVID-19. In the stage 2 experiment we will use the outcomes defined *a priori* in stage 1 to test whether the use of remdesivir use during acute COVID-19 is associated with subsequent development of Long COVID. In this protocol manuscript, we present the methods and results for stage 1, in which we *a priori* define Long COVID outcomes. In the discussion, we detail the experimental plan for the stage 2, where we will test the association of remdesivir use during acute COVID-19 and the association with subsequent development of Long COVID.

## Methods

### Study design, objectives and hypotheses

This is a retrospective cohort study using clinical data derived from electronic health records (EHR) performed in two stages. In stage 1, we *a priori* define the primary and secondary outcomes of Long COVID for use in stage 2. In stage 2, we will estimate the effect of remdesivir given during hospitalization for acute COVID-19 and the association with subsequent development of Long COVID.

*Stage 1*: The primary objective is to *a priori* define the outcomes of Long COVID. Here, we developed a cohort of hospitalized persons with and without documented acute COVID-19. The study cohort includes individuals hospitalized with a first episode of acute COVID-19 and the control cohort includes persons hospitalized without any current or history of COVID-19. We reasoned that features of Long COVID must be detectable as increased in the study cohort after hospitalization for acute COVID compared to the control cohort who were hospitalized for other reasons. The hypothesis for stage 1 is that features of Long COVID will be detectable in medically reported symptoms and conditions after hospitalization for acute COVID-19, compared to hospitalization for any other cause.

*Stage 2*: The primary objective is to test whether use of remdesivir during acute COVID-19 is associated with subsequent development of Long COVID. Here, we will limit analysis to the study cohort of persons hospitalized for acute COVID-19 and divide the cohort into exposed and unexposed groups with the exposure group defined by the receipt of remdesivir within 3 days of hospitalization. The hypothesis is that remdesivir use will be associated with decreased risk of developing Long COVID.

Stage 1 was completed prior to assessment of the exposure variable (receipt of remdesivir) in stage 2, thus the Long COVID outcomes were defined *a priori*; this study protocol manuscript memorializes the selection of Long COVID outcomes. In this protocol manuscript, the Methods and Results sections detail stage 1 processes and findings to *a priori* define Long COVID outcomes prior to testing the hypothesis of stage 2. The Discussion section contextualizes Stage 1 and presents further details of the experimental plan for Stage 2 to test whether use of remdesivir in acute COVID-19 is associated with subsequent development of Long COVID.

### Ethics statement

All procedures were reviewed and approved by the Providence Institutional Review Board (Providence IRB STUDY2023000049). Patient consent was waived because disclosure of protected health information for the study was determined to involve no more than a minimal risk to the privacy of individuals.

### Setting and participants

The study was conducted with data from Providence St Joseph Health (PSJH), an integrated health-care system that serves patients in 51 hospitals and 1085 clinics across seven states in the western United States. The timeframe analyzed for cohort selection for this study is from 05/01/2020 to 09/30/2022.

#### Study Cohort

Adults hospitalized for the first episode of acute COVID-19 during the study timeframe were identified who had an encounter in the three years prior to start of the study timeframe. Out of those patients with at least 1 encounter in the 3 years prior to study (05/01/2017 to 04/30/2020) we selected those who were hospitalized from 05/01/2020 to 09/30/2022 and who tested positive for the first time using SARS-CoV-2 nucleic acid amplification test (NAAT) or antigen test or had the first encounter diagnosis of COVID-19. We required that the patients were hospitalized 5 days before or up to 30 days after the positive test/diagnosis of COVID-19. Index date in the study cohort is defined as the earliest of: date of hospital admission, date of first positive SARS-CoV-2 test, or date of first COVID-19 encounter diagnosis. Patients were excluded who had received COVID-19 antiviral or antibody treatment within 30 days of the index date (**Table S1**), those enrolled in clinical trials for COVID-19, and those with prior evidence of SARS-CoV-2 infection (i.e. experiencing reinfection).

#### Control Cohort

Adults hospitalized for reasons other than COVID-19 during the same study timeframe who had no prior evidence of SARS-CoV-2 infection were identified who had an encounter in the three years prior to start of the study timeframe. Out of those patients with at least 1 encounter in the 3 years prior to study (05/01/2017 to 04/30/2020) we selected those who were hospitalized from 05/01/2020 to 09/30/2022 and who had no records of a positive test of SARS-CoV-2 NAAT or antigen test and had no record of encounter diagnosis of COVID-19. Index date in the study cohort is defined as the hospital admission date. Patients were excluded who had received COVID-19 antiviral or antibody treatment within 30 days of the index date.

#### Negative Exposure Cohort

To assess for spurious associations in the Long COVID outcomes selected, we tested a separate cohort with a random exposure not expected to have any association with the outcomes. The random exposure was receipt of influenza vaccine on even versus odd (referent category) days of the month. We assessed a two-year period prior to the COVID-19 pandemic. Out of those patients with at least 1 encounter in the 3 years prior to the negative exposure cohort time frame (05/01/2014 to 04/30/2017) we selected those who received influenza vaccine in the time frame of 05/01/2017 to 04/30/2019. Index date is defined as the receipt of influenza vaccine date. In this cohort, there were 1,122,878 unique individuals. Of these, 532,320 received influenza vaccine on an even calendar day and 590,558 received influenza vaccine on an odd calendar day.

### Participant Timeline

Participants for the study and control cohorts were hospitalized between 05/01/2020 to 09/30/2022. Timelines for individual participants were constructed with baseline, acute and post-acute periods (**Figure S1**). The index date (time zero, T_0_) for the study cohort (hospitalized with COVID) is defined as the hospital admission date, the first positive test date for SARS-CoV-2, or the first encounter diagnosis date for COVID-19, whichever came first. T_0_ for the control cohort (hospitalized without COVID) is defined as the hospital admission date. The acute period is defined from T_0_ to T_0_+89 days. Measurement of outcomes starts at the post-acute period, starting from T_0_+90 days to T_0_+365 days. To study incident outcomes, we assessed for history of the individual outcome in the baseline period, which differed for outcomes which are acute or transient symptoms in nature (e.g. cough) or chronic conditions (e.g. heart failure). The designations of symptoms or conditions as acute or chronic are given in supplemental **Table S2**. For chronic conditions, the baseline period is defined as any time prior to T_0_. For acute symptoms, the baseline period is defined as within 365 days before T_0_ (**Figure S1**).

### Outcomes

#### Long COVID Outcomes

The primary outcome is an incident “Long COVID” detected between 90 and *365* days after hospitalization. Long COVID definitions were aligned with an emerging literature consensus,^9,12,13^ especially from NASEM.^2^ The primary outcome was a combined outcome of ICD-10 code U09.9 (Post-COVID Conditions) or newly diagnosed individual symptoms or conditions which were differential in our study cohort after hospitalization for acute COVID-19 compared to the control cohort after hospitalization without COVID-19. Secondary outcomes included each of the individual components of the combined primary outcome (except U09.9). To select the incident components, we started with the 821 high dimensional outcomes (379 diagnoses, 380 medication classes and 62 laboratory abnormalities) first studied by Al-Aly^1^ and then narrowed to a shorter list of 27 final outcomes, similar to what has been done in this group’s subsequent work^3^. Expert opinions from 2 authors (JDG, JJH) were used to select individual Long COVID outcomes from specific diagnosis codes consistent with evolving definitions of Long COVID and emerging literature,^2,9,12,13^ and understanding of pathophysiology.^5,6^ Individual diagnosed symptoms or conditions were based on ICD-10 codes and laboratory or other biomarkers (i.e. creatinine, body mass index, etc). ICD-10 codes were included if they represented the syndromes of Long COVID and if they were not clearly related to another cause. Specifically, alternative explanations and codes for procedural complications were removed, but codes relating to the conditions in pregnancy or other conditions which could be exacerbated by Long COVID were retained. The selection process was done iteratively, with the final outcome selection accommodating the ability to detect these Long COVID outcomes in the context of incident diagnoses or biomarkers in the follow-up post-acute period in the study versus control cohorts. We reasoned that individual manifestations of Long COVID should be more common in a person hospitalized with COVID-19, compared to those hospitalized for other reasons. Thus, the ability to detect individual Long COVID manifestations in our context was a pretext for selection of the individual Long COVID manifestations as a secondary outcome (and thus a component of the combined primary outcome). Incident symptoms and conditions which were newly diagnosed since the index hospitalization that did not occur in a relevant baseline period were selected. At least 100 total incident events were required to have occurred in the follow-up period in either the study or control cohorts to be selected as a pre-specified outcome. While selection of incident symptoms and conditions do not allow for the study of worsening of pre-existing conditions,^14^ we did not feel that worsening could be adequately assessed in these observational data. ICD-10 codes and biomarkers with thresholds for the secondary outcomes which are components of the combined primary outcome are given in supplemental **Table S2**. Exploratory outcomes are the original set of high dimensional outcomes presented by Al-Aly (**Tables S3 - S5**).^4^

#### Negative Outcomes

Conditions with no suspected relationship to COVID-19 or its treatment were studied as negative outcomes, to increase confidence for inference that detected long COVID outcomes are not spurious. Negative outcomes included neoplasms (benign or malignant) and hernias (**Table S2**). The negative outcomes are not thought to be associated with acute COVID or long COVID and are expected not to differ in follow-up assessment between the study and control cohorts.

### Covariates

#### Pre-specified covariates

included those variables which were defined in advance as standard risk factors for poor outcomes in acute COVID-19 or those variables possibly associated with the propensity to receive the antiviral remdesivir including: age, sex, race, ethnicity, commercial insurance, Charlson Comorbidity Index (CCI),^15,16^ history of kidney disease or eGFR < 60 mL/min/1.73m2, history of liver disease or elevated liver function tests (ALT or AST), WHO ordinal scale score (OSS, as moderate OSS 3, severe OSS 4-5, critical OSS 6-7) which indicates supplemental oxygen use including mechanical ventillation,^17^ fully vaccinated for COVID-19,^18^ time from last COVID-19 vaccination to T_0_, SARS-CoV-2 variant era (early: 03/01/2020 - 06/25/2021, delta 06/26/2021 - 12/24/2021, omicron after 12/25/2021), number of encounters in year prior to T_0_, systemic steroid use during acute COVID-19 (in prednisone equivalents),^19^ baricitinib or tocilizumab use during acute COVID-19, and immunocompromised status.^20^ Age in years was recorded on T_0_. CCI was computed using all comorbidity data prior to T_0_, but age was removed from the score as we separately controlled for age. Number of encounters and commercial insurance were extracted from data available from a year prior to T_0_. We extracted steroid quantity, WHO score and Baricitinib/Tocilizumab covariates for the acute period starting T_0_ to T_0_+30 days. Missing values for race, ethnicity, and commercial insurance were replaced as ‘Other’. Additional coding elements for relevant pre-specified covariates is given in supplemental **Table S6**.

#### High-dimensional covariates

821 covariates were selected from data domains including diagnoses, medication use, and laboratory tests as previously described.^4^ Clinical Classifications Software Refined (version 2023.1) was used to classify ICD-10 codes into 379 diagnostic categories (**Table S3**). The VA drug classification system was used to classify RxNorm into 380 medication classes (**Table S4**). As previously defined in Al-Aly et al., we categorized 62 laboratory abnormalities from 38 laboratory tests (**Table S5**).[6] High dimensional covariates were assessed in the 365 days prior toT0. Missingness of these variables are assumed normal (i.e. no condition, med or lab abnormality present).

### Statistical Analysis

The analysis is conducted with a three-step workflow (**Figure S2**), as previously described.^4^ First, for each individual secondary outcome, an outcome-specific cohort was constructed removing participants with the outcome discovered in the relevant baseline timeframe. Next, for each outcome-specific cohort, overlap weights were computed from covariates and the populations balanced using overlap weights. Finally, outcomes were computed including the hazard ratio, incidence rate and excess burden. For the assessment of the combined primary outcome, all patients were included, and components of the combined primary outcome were counted only if the component outcome did not occur in the relevant baseline period.

#### Covariate Balance using Overlap Weights

The study and control cohorts were balanced using overlap weights based on propensity scores for variables which may confound the examined associations. Pre-specified baseline covariates including age, sex, race, ethnicity, CCI, commercial insurance, number of encounters in the year prior to index, history of liver disease or elevated liver function tests, kidney dysfunction, immunocompromised status, time since last COVID-19 vaccine, fully vaccinated indicator, variant era of admission, and variables based on the index hospital admission steroid quantity (in mg of prednisone equivalents), highest WHO score, and receipt of baricitinib or tocilizumab. The set of high dimensional covariates were considered for inclusion in the model if the event occurred in the baseline period in ≥ 100 subjects in each group. We then calculated univariate relative risk for each high dimensional covariate, differential in the baseline period between study and control cohorts. The top 100 high dimensional covariates with the highest relative risk in univariate analysis and all pre-specified covariates were included in the propensity score logistic model. Group membership is coded as Y=1 for the study cohort (hospitalized with acute COVID-19) and Y=0 for the control cohort (hospitalized without COVID-19). We defined propensity score as Pr(Y=1|X), or the probability of membership in the study cohort given the observed characteristics, or covariates, denoted by X. Propensity scores are computed using logistic regression, where group membership is the dependent variable and the covariates are the set defined by pre-specified along with the top 100 high-dimensional covariates. Overlap weights are defined as 1 – Pr(Y=1|X) for the study group and Pr(Y=1|X) for the control group. Covariate balance after weighting is assessed by calculating the standardized mean difference (SMD), using a threshold of SMD < 0.1 to indicate good balance.

#### Hazard Ratio, Incidence Rates and Excess Burden

Incident outcomes were counted and presented as raw data. After balancing, incidence rates for each outcome per 1000 persons at 360 days (end of follow-up) are computed as 1000 times the number of incident outcomes divided by person-time, where person-time is the sum of the total number of days in the follow up period for the outcome-specific cohort divided by 365 to express in person years. Excess burden for each outcome-specific cohort is calculated as the difference of incidence rates between exposed and unexposed groups. For the combined primary outcome on the entire cohort and for individual secondary outcomes for each outcome-specific cohort, hazard ratio (HR) is calculated for each outcome-specific cohort using Cox proportional hazard model using the overlap weighted populations. Robust standard errors computed using the Huber sandwich estimator were used to derive the 95% confidence intervals (95%CIs). Proportional hazard assumptions were tested for all outcomes with Schoenfeld residuals. The HR was not constant during the follow-up period for all secondary outcomes, thus the HR should be interpreted as the weighted average of the potentially time-varying HR for secondary outcomes.^21^ Only outcomes with at least 10 events in each group were modeled in the exploratory analyses (high dimensional) and only outcomes with at least 10 outcomes in each group and 100 total outcomes were considered for selection as a pre-specified secondary outcome. For the combined primary outcome, the event occurs when a diagnosis of Post-Covid Condition (U09.9) or an incident pre-specified outcome occurs, and persons are censored at the first such event. No persons in the cohort are excluded from this analysis of the combined primary outcome, though events are only considered as incident if the condition did not exist in the relevant baseline period (see Participant Timeline, above). Deaths which occurred during the acute period (T_0_ to T_0_ + 89 days) did not allow for contribution of person-time to the primary combined outcome as the follow-up period for outcomes assessment started at T_0_ + 90 days. Repeat COVID diagnoses or labs in the acute period were considered as part of the same event and were not considered reinfections. Death and reinfection after T_0_ + 90 days have been treated as competing risks and we have right-censoring at these events to calculate the cause-specific hazard model. Individuals were censored when an outcome (primary or secondary) occurred, or were right censored at the earliest of death, reinfection, last known alive date, or 365 days, whichever occurred first. In the alternative combined outcome, death after T_0_ is considered an event counted towards the combined primary outcome, instead of right censored.

#### Inference for Multiple Comparisons

In stage 1 of this protocol, we first test the combined primary and individual component secondary outcomes in the study versus control cohorts (hospitalized with vs. without COVID). For the combined primary outcome p-value < 0.05 is considered statistically significant. For secondary outcomes (individual components of the primary outcome), the Holm-Bonferroni correction is applied to address the multiple testing problem. A statistically significant p-value for each of the individual outcomes are p1 < α/*m*, p2 < α/(*m*-1), p3 < α/(*m*-2), …, pm < α, where *m* is the number of statistical tests and α = 0.05 and the p-values are ordered from lowest to highest. If any test fails to reject the null hypothesis, all further tests fail. Holm-Bonferroni caps the maximum family-wise error rate for all statistical tests to α = 0.05. We next test the same panel of outcomes on the negative exposure cohort (influenza vaccine on even or odd calendar days).

#### Down-sampling Experiment

In stage 1, we *a priori* select the Long COVID combined primary and secondary outcomes by testing associations of the outcomes in the study cohort (hospitalized with COVID) versus control cohort (hospitalized without COVID). Anticipating a much smaller sample size for the stage 2 experiment (the study cohort hospitalized with COVID is subset to those with and without remdesivir exposure), we perform a set of down-sampling experiments to anticipate how stable the assessment of the Long COVID combined primary and individual secondary outcomes would be in a simulated smaller cohort. To check the robustness of the pre-specified outcomes in a simulated smaller cohort, we sample without replacement for the anticipated size of the remdesivir vs. no remdesivir cohorts. In N = 100 bootstrap samples, we then run the entire analysis pipeline, as above, and calculate the mean (SD) and 95%CI for the HR and the mean (SD) for the p-value across the 100 replicates. We then calculate the anticipated proportion of bootstrapped samples which would maintain the positive associations given the smaller sample size using two false discovery rate (FDR) corrections, the Benjamini-Hochberg (FDR-BH)^22^ and the Benjamini-Yekutieli (FDR-BY)^23^ procedures.

The lifelines library (Python) is used for building Cox proportional hazards models and statsmodels (Python) is used for statistical analyses ^6^.^24^

## Results

### Study and Control Cohorts

Between 05/01/2017 - 04/30/2020, there were 10,164,480 unique individuals with ≥ 1 clinical encounter. Of these, 573,007 persons were hospitalized between 05/01/2020 - 09/30/2022. There were 128,052 persons hospitalized with a positive SARS-CoV-2 test or encounter diagnosis of COVID-19, and 445,030 persons with no record of a positive SARS-CoV-2 test and no encounter diagnosis of COVID-19, at time of or prior to hospitalization. Of those hospitalized with COVID, the study cohort included 45,540 adults (1) hospitalized 5 days before or up to 30 days after the positive SARS-CoV-2 test or encounter diagnosis of COVID-19; (2) No other COVID-19 antiviral or antibody treatments other than remdesivir within ±30 days of T_0_; (3) No remdesivir /placebo in clinical trial; and (4) were alive at T_0_ + 90 days and evaluable for the combined primary outcome. Of those hospitalized without COVID, the control cohort included 409,186 adults (1) without any COVID-19 antiviral or antibody treatment within ±30 days of T_0_ and (2) Alive at T_0_ + 90 days (**Figure 1**). For the cohort of hospitalized with COVID, 8,147 (17.9%) were diagnosed with a diagnosis code only, 11,588 (25.4%) were diagnosed with a lab only and 28,577 (62.7%) were diagnosed with both.

**Figure 1:**
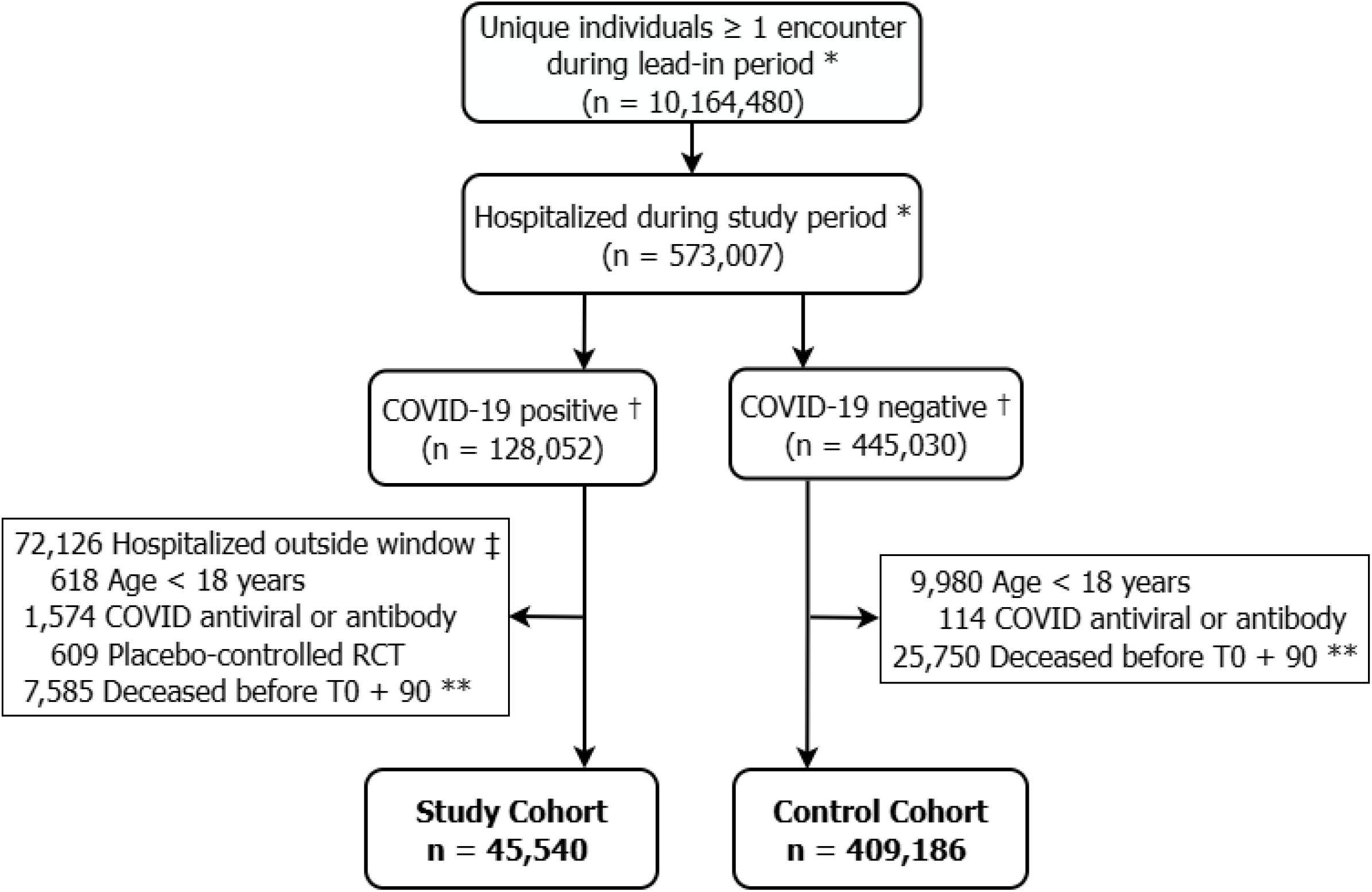
Cohorts Flow Diagram for Study Cohort (hospitalized with COVID) and Control Cohort (hospitalized without COVID). ^*^ Lead-in period = 05/01/2017 to 04/30/2020; Study period = 05/01/2020 to 09/30/2022; † Defined by positive SARS-CoV-2 Ag or NAT test or COVID-19 diagnosis (COVID-19 positive) vs. no record of these now or in the past (COVID-19 negative). ^‡^ Window for COVID hospitalization was admission date ≤ 5 days before or ≤ 30 after SARS-CoV-2 positive test or COVID-19 diagnosis. ^* *^Time zero (T0) was defined as the earliest of: hospital admission date, first positive SARS-CoV-2 test date, or first COVID-19 encounter diagnosis date.

**Figure 2:**
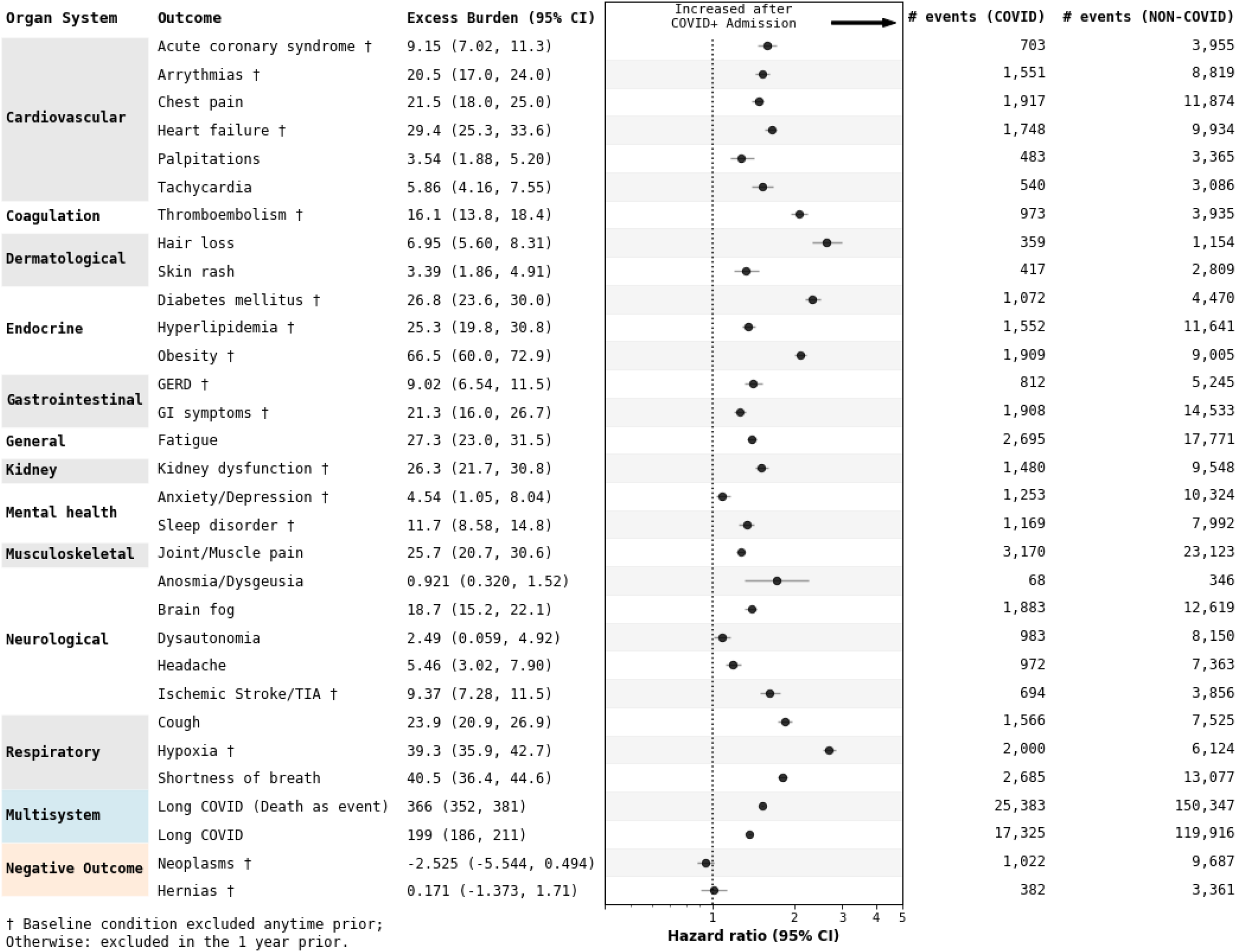
Forest Plot for the combined primary and secondary Long COVID outcomes in the study cohort (hospitalized with COVID) vs. control cohort (hospitalized without COVID).

Baseline characteristics of the included cohort are given in **Table 1**. Age was mean (standard deviation, SD) was 60.7 (19.3) vs. 57.3 (21.3) years and age 46% and 37% were male in the study vs. control cohorts respectively. The majority of persons in both cohorts were Caucasian, and non-Hispanic. The mean (SD) CCI was 1.27 (1.88) vs. 1.03 (1.69) in the study vs. control cohorts. Higher CCI reflected an increased baseline prevalence of common comorbidities in the study cohort such as obesity (49.2% vs. 44.8%), hyperlipidemia (46.0% vs. 39.3%), diabetes mellitus (26.5% vs. 20.2%), heart failure (25.7% vs. 22.8%) and kidney dysfunction (37.0% vs. 32.4%), in the study vs. control cohorts respectively. Anxiety/depression were similar between the cohorts (77%). In the control cohort, a higher proportion had commercial insurance and there were more encounters in the baseline period. In the study cohort, a higher proportion had a history of liver disease /elevated liver function tests or kidney dysfunction, were immunocompromised and were more severe based on the WHO score. 28.8% of those hospitalized for reasons other than COVID were fully vaccinated, compared to 23.6% in the cohort hospitalized for COVID. Corticosteroid and baricitinib/tocilizumab use during the index hospitalization was much higher in the study cohort, consistent with use to treat COVID-19 during the index hospitalization for acute COVID. A lower proportion of hospitalizations occurred during the origin variant era and higher proportion during the Delta and Omicron variant eras in the study vs. control cohorts. Relative risk was assessed for each of the high dimensional covariates, and the top 100 were selected for inclusion in the weighting by descending relative risk (**Table S7**). Applying the overlap weights based on the propensity score from the pre-specified covariates and the top 100 high dimensional covariates normalized baseline variables including age, sex, race, ethnicity, insurance, vaccination, variant era, WHO ordinal scale score, steroid use, immunocompromised status and 100 clinical features with SMD ≤ 0.01 for all baseline variables (**Table 1, Table S8**).

**Table 1:** Balance of Baseline Covariates before and after overlap weighting in the study cohort (hospitalized with COVID) and control cohort (hospitalized without COVID).

|  | Population before overlap weights |  | Population after overlap weights |  |  |
| --- | --- | --- | --- | --- | --- |
| Feature * | Study | Control | Study | Control | SMD |
| Age † | 60.7 (19.3) | 57.2 (21.3) | 60.3 (19.7) | 60.3 (20.6) | 2.38E-04 |
| Sex | 21021 (46.2%) | 150413 (36.8%) | 21021 (44.5%) | 150413 (44.5%) | 9.25E-04 |
| Race |  |  |  |  |  |
| Black | 1730 (3.80%) | 14316 (3.50%) | 1730 (3.84%) | 14316 (3.88%) | 2.20E-03 |
| Other | 12020 (26.4%) | 88846 (21.7%) | 12020 (25.6%) | 88846 (25.5%) | 1.35E-03 |
| White | 31790 (69.8%) | 306024 (74.8%) | 31790 (70.6%) | 306024 (70.6%) | 3.59E-04 |
| Ethnicity |  |  |  |  |  |
| Hispanic | 8888 (19.5%) | 50333 (12.3%) | 8888 (18.3%) | 50333 (18.3%) | 3.09E-04 |
| Not Hispanic | 31049 (68.2%) | 307004 (75.0%) | 31049 (68.9%) | 307004 (68.9%) | 4.69E-04 |
| Other | 5603 (12.3%) | 51849 (12.7%) | 5603 (12.8%) | 51849 (12.8%) | 2.91E-04 |
| Charlson Comorbidity Index † | 1.27 (1.88) | 1.03 (1.69) | 1.23 (1.85) | 1.23 (1.81) | 8.15E-04 |
| Commercial Insurance | 9506 (20.9%) | 114484 (28.0%) | 9506 (21.0%) | 114484 (21.0%) | 4.09E-05 |
| Number encounters † | 0.541 (1.49) | 0.758 (1.66) | 0.564 (1.56) | 0.563 (1.07) | 1.10E-03 |
| Liver disease | 2026 (4.45%) | 13246 (3.24%) | 2026 (4.17%) | 13246 (4.15%) | 1.49E-03 |
| Kidney disease | 10097 (22.2%) | 63614 (15.5%) | 10097 (20.9%) | 63614 (21.0%) | 1.08E-03 |
| Immunocompromised | 8600 (18.9%) | 49305 (12.0%) | 8600 (17.3%) | 49305 (17.4%) | 2.77E-03 |
| Fully vaccinated | 10752 (23.6%) | 117715 (28.8%) | 10752 (24.6%) | 117715 (24.6%) | 3.16E-04 |
| Time since last COVID-19 vaccine † | 794 (375) | 734 (415) | 784 (381) | 784 (385) | 1.41E-04 |
| Baricitinib/Tocilizumab | 1400 (3.07%) | 8 (0.00196%) | 1400 (0.0571%) | 8 (0.021%) | 1.83E-02 |
| Steroid quantity † | 154 (347) | 36.8 (158) | 104 (201) | 104 (382) | 6.40E-04 |
| WHO score |  |  |  |  |  |
| Critical | 8415 (18.5%) | 31308 (7.65%) | 8415 (13.8%) | 31308 (13.7%) | 2.54E-03 |
| severe | 18303 (40.2%) | 144077 (35.2%) | 18303 (40.4%) | 144077 (40.4%) | 5.67E-04 |
| Non severe | 18822 (41.3%) | 233801 (57.1%) | 18822 (45.9%) | 233801 (46%) | 1.20E-03 |
| Variant era |  |  |  |  |  |
| Early | 19328 (42.4%) | 235799 (57.6%) | 19328 (45.8%) | 235799 (45.8%) | 1.03E-04 |
| Delta | 10639 (23.4%) | 75511 (18.5%) | 10639 (20.9%) | 75511 (20.9%) | 1.25E-03 |
| Omicron | 15573 (34.2%) | 97876 (23.9%) | 15573 (33.2%) | 97876 (33.3%) | 1.19E-03 |

Abbreviations: SMD: standard mean difference; SD: standard deviation; WHO score: World Health Organization, ordinal scale score.
\* Timing of all features is given in methods. Time since last COVID-19 vaccine in days. Steroid quantity is given in mg of prednisone equivalents. WHO score is moderate if 3, severe if 4-5 and critical if 6-7. Variant eras: Early: 03/01/2020 - 06/25/2021, Delta 06/26/2021 - 12/24/2021, Omicron after 12/25/2021.
† Data is given as mean (SD) as indicated; otherwise, data given as count (%).

### Primary, Secondary and Exploratory Outcomes

#### Primary Outcomes

The combined primary Long Covid outcome occurred in 17,325 of 45,540 (38.0%) persons and 119,916 of 409,186 (29.3%) persons in the study cohort (hospitalized with COVID) and control cohort (hospitalized without COVID), respectively. The unweighted incidence rate was 733 per 1,000 person years (95%CI: 722, 744) in the study cohort and 521 per 1,000 persons years (95%CI: 518, 524) for unadjusted excess burden in the study cohort of 212 per 1,000 person years (95%CI: 201, 224). The weighted incidence rate was 715 per 1,000 person years (95%CI: 703, 727) in the study cohort and 516 per 1,000 person years (95%CI: 513, 519) in the control cohort for adjusted excess burden in the study cohort of 199 per 1,000 person years (95%CI: 186, 211). The hazard ratio for the Long COVID combined primary outcome was 1.37 (95%CI: 1.35, 1.40), p < 0.0001. This corresponds to a 37% increased risk in the study compared to control cohorts of having Post-COVID Conditions (U09.9) diagnosis or ≥ 1 of the individual secondary outcomes in the follow-up period. Diagnosis with U09.9 occurred in 689 (1.51%) and 163 (0.040%) persons were in the study and control cohorts, respectively. The weighted excess burden of U09.9 diagnosis was 16.5 per 1,000 persons (95%CI: 14.9, 18.2) in the study cohort and HR was 29.2 (95%CI: 24.5, 34.9) for having U09.9 diagnosis, or 29 times increased risk for U09.9 in the study cohort (**Table S9**). Death occurred in 9,073 persons in the study cohort (hospitalized with COVID), 7,585 of which occurred prior to 90 days, and in 39,293 persons in the control cohort (hospitalized without COVID), 25,750 of which occurred prior to 90 days. In the alternative combined outcome (where death is treated as an event instead of a competing risk), the HR for Long COVID combined primary outcome was 1.53 (95%CI 1.51, 1.55), p < 0.0001 or weighted excess burden of 366 (95%CI: 352, 381) in the study cohort (**Table 2** and **Table S9**).

**Table 2:** Hazard Ratios assessing association of the combined primary outcome (Long COVID) and 27 secondary outcomes (individual components of the combined primary outcome) in the study cohort (hospitalized with COVID) and control cohort (hospitalized without COVID).

| Organ | Diagnosis | HR (95%CI) | p-value |
| --- | --- | --- | --- |
| **Primary Combined Outcome** |  |  |  |
| Multisystem | Long COVID<br>(Death as competing risk) | 1.37 (1.35, 1.40) | < 0.0001 |
| **Alternative Combined Outcome** |  |  |  |
| Multisystem | Long COVID<br>(Death as event) | 1.53 (1.51, 1.55) | < 0.0001 |
| **Secondary Outcomes** |  |  |  |
| Cardiovascular | Acute coronary syndrome † | 1.59 (1.47, 1.73) | < 0.0001 |
|  | Arrhythmias † | 1.53 (1.44, 1.62) | < 0.0001 |
|  | Chest pain | 1.48 (1.40, 1.55) | < 0.0001 |
|  | Heart failure † | 1.65 (1.56, 1.73) | < 0.0001 |
|  | Palpitations | 1.28 (1.16, 1.42) | < 0.0001 |
|  | Tachycardia | 1.53 (1.39, 1.68) | < 0.0001 |
| Coagulation | Thromboembolism † | 2.09 (1.94, 2.25) | < 0.0001 |
| Dermatological | Hair loss | 2.64 (2.33, 2.99) | < 0.0001 |
|  | Skin rash | 1.33 (1.20, 1.48) | < 0.0001 |
| Endocrine | Diabetes mellitus † | 2.34 (2.19, 2.51) | < 0.0001 |
|  | Hyperlipidemia † | 1.36 (1.29, 1.44) | < 0.0001 |
|  | Obesity † | 2.10 (2.00, 2.21) | < 0.0001 |
| Gastrointestinal | GERD † | 1.41 (1.31, 1.53) | < 0.0001 |
|  | GI Symptoms † | 1.26 (1.20, 1.33) | < 0.0001 |
| General | Fatigue | 1.40 (1.34, 1.46) | < 0.0001 |
| Kidney | Kidney dysfunction † | 1.52 (1.44, 1.61) | < 0.0001 |
| Mental health | Anxiety / Depression † | 1.09 (1.03, 1.16) | 0.0045 |
|  | Sleep disorder † | 1.34 (1.25, 1.42) | < 0.0001 |
| Musculoskeletal | Joint/Muscle pain | 1.27 (1.22, 1.31) | < 0.0001 |
| Neurological | Brain fog | 1.39 (1.32, 1.46) | < 0.0001 |
|  | Dysautonomia | 1.08 (1.01, 1.16) | 0.024 |
|  | Headache | 1.19 (1.12, 1.28) | < 0.0001 |
|  | Ischemic Stroke/TIA † | 1.63 (1.50, 1.77) | < 0.0001 |
|  | Smell/Taste problems | 1.73 (1.32, 2.27) | < 0.0001 |
| Respiratory | Cough | 1.85 (1.75, 1.96) | < 0.0001 |
|  | Hypoxia † | 2.69 (2.55, 2.84) | < 0.0001 |
|  | Shortness of breath | 1.81 (1.74, 1.89) | < 0.0001 |
| **Negative Outcomes** |  |  |  |
| Neoplasms | Neoplasms† | 0.94 (0.88, 1.01) | 0.082 |
| Hernias | Hernias† | 1.01 (0.908, 1.13) | 0.81 |

† : Baseline condition excluded anytime prior to T0, otherwise excluded in the 1 year prior to T0.

#### Secondary Outcomes

Among the secondary outcomes (individual components of the combined primary outcome), raw events counts were common in both the study and control cohorts. All events occurred in >500 persons in the study cohort and in >1,000 persons in the control cohort, except for anosmia/dysgeusia, palpitations, skin rash & hair loss (**Table S9**). All 27 secondary outcomes show positive associations in the study compared to control cohorts. Strong associations were seen for the following incident diagnoses: hypoxia HR 2.69 (95%CI: 2.55, 2.84), hair loss HR 2.64 (95%CI: 2.33, 2.99), diabetes mellitus HR 2.34 (95%CI: 2.19, 2.51), thromboembolism HR 2.09 (95%CI: 1.94, 2.25), obesity HR 2.10 (95%CI: 2.00, 2.21), p < 0.0001 for each of these outcomes. Some outcomes had weaker detected associations such as palpitations HR 1.28 (95%CI: 1.16, 1.42), skin rash HR 1.33 (95%CI: 1.20, 1.48), hyperlipidemia HR 1.36 (1.29, 1.44), gastrointestinal symptoms HR 1.26 (95%CI: 1.20, 1.33), sleep disorder HR 1.34 (95%CI: 1.25, 1.42), joint/muscle pain HR 1.27 (95%CI: 1.22, 1.31), headache HR 1.19 (95%CI: 1.12, 1.28), anxiety/depression HR 1.09 (95%CI: 1.03, 1.16), p=0.0045, and dysautonomia HR 1.08 (95%CI: 1.01, 1.16), p=0.024. After adjustment for multiplicity with the Holm-Bonferroni correction, all 27 secondary outcomes were associated with the study cohort (hospitalized with COVID) and met the threshold for statistical significance at p < 0.0001, except as indicated above (**Table 2**). Anosmia/dysgeusia was associated with prior COVID hospitalization, HR 1.73 (95%CI: 1.32, 2.27), p < 0.0001, but wider confidence intervals reflected fewer recorded diagnoses. Among the negative outcomes tested, neither neoplasms (p = 0.082) nor hernias (p = 0.81) were statistically significant between the study and control cohorts (**Table 2**).

#### Exploratory Outcomes

Of the 821 exploratory outcomes, 742 were evaluable, and 279 outcomes (37.6%) passed the Holm-Bonferroni correction for multiple hypothesis testing (**Table S10**). Of these, 96% had positive associations with hospitalization for COVID. Many of these outcomes were noted as associated with known post-acute sequelae after covid such as diseases of the respiratory system, e.g. pneumonia HR 2.90 (95%CI 2.76, 3.05), or respiratory tract medications, e.g. antitussives/expectorants HR 1.96 (95%CI 1.85, 2.07) or bronchodilators HR 1.63 (95%CI 1.56, 1.69). Other conditions noted below the adjusted level of significance with stronger associations (HR > 1.5) included conditions which were encompassed by the secondary outcomes. These included diagnoses and medications associated with diabetes, hyperlipidemia, obesity, renal dysfunction, acute pulmonary embolism. Other notable associations include immunity disorders and use of immunoglobulins, organ transplant status, and additional use of antimicrobials (e.g. fourth generation cephalosporins and anthelminthics), and vitamins and dietary supplements (e.g. vitamin C and zinc). Associations noted in these exploratory outcomes should be hypothesis generating only and require confirmation or further exploration. In comparison, in the negative exposure cohort (influenza vaccination on even versus odd days of the month), of 821 exploratory, 759 were evaluable and only 1 outcome (0.13%) passed the Holm-Bonferroni correction for multiple testing (**Table S11**).

### Down-sampling Experiment

To test the robustness of the associations of primary and secondary outcomes to a smaller sample size, we first assessed the proportion of the study cohort (hospitalized with COVID) who did and did not receive remdesivir. In the study cohort, 14,866 of 45,540 persons (32.6%) received remdesivir within 3 days of hospital admission, 633 (1.4%) received remdesivir after 3 days of hospitalization and 30,041 (65.9%) did not receive any remdesivir. We then sampled without replacement for n=14,866 randomly selected persons down-sampled from the study cohort, and n = 30,041 randomly selected persons down-sampled from the control cohort, and calculated the HR for each of the pre-specified secondary outcomes. We repeated N=100 times for the bootstrapped random down-samples. The mean HR in the down-sampled cohorts across the bootstrapped cohorts are similar across all secondary outcomes to the main cohort. The same five outcomes had strong associations (mean HR > 2): hypoxemia, hair loss, diabetes mellitus, thromboembolism, and obesity. Smell and taste disorder also had strong association with the mean HR (SD) = 2.00 (0.583), but a wide confidence interval (95%CI: 1.01, 3.25). The FDR-adjusted p-value was < 0.05 by both FDR-BH and FDR-BY methods in ≥ 93% of bootstrapped samples for 21 of 27 pre-specified secondary outcomes. However, significant imprecision would be expected by a lower proportion of FDR-adjusted p-value for palpitations, skin rash, anxiety/depression, dysautonomia, headache, and anosmia/dysgeusia. The most poorly performing secondary outcomes demonstrating the lowest proportion of statistically significant associations in the expected FDR-adjusted p-values are for dysautonomia with 15% and 7%, anxiety/depression with 16% and 9%, by FDR-BH and FDR-BY corrections, respectively. For the negative outcomes of hernias and neoplasms, ≤ 7% of expected FDR-adjusted p-values would be significant at the α < 0.05 level (**Table 3** and **Table S12**). These results suggest overall excellent expected performance in most of the *a priori* selected outcomes when used in a smaller dataset anticipated for stage 2, with marginal expected performance in a few of the secondary outcomes.

**Table 3:** Results of down-sampling experiment with N=100 replicates.

| Organ System | Diagnosis | HR, mean<br>(95%CI) | p-value,<br>mean | FDR, BH<br>(%) * | FDR, BY<br>(%) * |
| --- | --- | --- | --- | --- | --- |
| **Secondary Outcomes** |  |  |  |  |  |
| Cardiovascular | Acute coronary syndrome † | 1.60 (1.41, 1.83) | 4.30E-05 | 100% | 100% |
|  | Arrhythmias † | 1.50 (1.35, 1.66) | 7.36E-07 | 100% | 100% |
|  | Chest pain | 1.46 (1.35, 1.62) | 2.24E-08 | 100% | 100% |
|  | Heart failure † | 1.62 (1.48, 1.77) | 1.09E-11 | 100% | 100% |
|  | Palpitations | 1.29 (1.08, 1.53) | 8.62E-02 | 66% | 46% |
|  | Tachycardia | 1.53 (1.26, 1.87) | 7.47E-03 | 98% | 93% |
| Coagulation | Thromboembolism † | 2.04 (1.72, 2.32) | 1.12E-09 | 100% | 100% |
| Dermatological | Hair loss | 2.58 (1.99, 3.31) | 8.27E-06 | 100% | 100% |
|  | Skin rash | 1.35 (1.12, 1.61) | 6.29E-02 | 73% | 53% |
| Endocrine | Diabetes mellitus † | 2.36 (2.05, 2.66) | 3.64E-19 | 100% | 100% |
|  | Hyperlipidemia † | 1.36 (1.21, 1.50) | 9.11E-05 | 100% | 100% |
|  | Obesity † | 2.14 (1.92, 2.31) | 6.74E-28 | 100% | 100% |
| Gastrointestinal | GERD † | 1.42 (1.26, 1.61) | 2.01E-03 | 99% | 99% |
|  | GI Symptoms † | 1.28 (1.19, 1.42) | 5.92E-04 | 100% | 99% |
| General | Fatigue | 1.38 (1.28, 1.48) | 4.41E-08 | 100% | 100% |
| Kidney | Kidney dysfunction † | 1.51 (1.33, 1.71) | 3.16E-07 | 100% | 100% |
| Mental health | Anxiety / Depression † | 1.08 (0.974, 1.22) | 3.23E-01 | 16% | 9% |
|  | Sleep disorder † | 1.35 (1.18, 1.55) | 1.92E-03 | 99% | 94% |
| Musculoskeletal | Joint/Muscle pain | 1.27 (1.18, 1.36) | 1.36E-05 | 100% | 100% |
| Neurological | Brain fog | 1.38 (1.26, 1.57) | 4.79E-06 | 100% | 100% |
|  | Dysautonomia | 1.08 (0.926, 1.24) | 3.68E-01 | 15% | 7% |
|  | Headache | 1.18 (1.05, 1.31) | 9.33E-02 | 55% | 33% |
|  | Ischemic Stroke/TIA † | 1.62 (1.35, 1.90) | 6.93E-04 | 99% | 99% |
|  | Smell/Taste problems | 2.00 (1.01, 3.25) | 1.17E-01 | 58% | 42% |
| Respiratory | Cough | 1.83 (1.67, 2.00) | 1.92E-15 | 100% | 100% |
|  | Hypoxia † | 2.52 (2.22, 2.84) | 2.93E-29 | 100% | 100% |
|  | Shortness of breath | 1.78 (1.62, 1.97) | 1.55E-21 | 100% | 100% |
| **Negative Outcomes** |  |  |  |  |  |
| Neoplasms | Neoplasms† | 0.94 (0.836, 1.08) | 4.04E-01 | 7% | 3% |
| Hernias | Hernias† | 1.02 (0.826, 1.24) | 5.66E-01 | 1% | 1% |

Abbreviations: GERD: Gastrointestinal Reflux Disease; GI: Gastrointestinal; TIA: Transient Ischemic Attack; HR (95%CI): Hazard Ratio (with 95% Confidence Intervals); FDR: False Discovery Rate; BH: Benjamini-Hochberg method; BY: Benjamini-Yekutieli method.
† : Baseline condition excluded any time prior to T0, otherwise excluded in the 1 year prior to T0.
\* FDR, BH or FDR, BY indicate the proportion of adjusted p-values < 0.05 in the 100 replicates by the FDR Benjamini-Hochberg or Benjamini-Yekutieli methods, respectively.

## Discussion

Long COVID is a newly described infection-associated chronic condition. The NASEM committee noted that the definition of Long COVID is broad and may need to be further specified for certain research purposes.^3,14^ Here in this protocol manuscript, we pre-specify Long COVID outcomes to be used to address a subsequent research question. We will use the definitions for Long COVID pre-specified here, and the analytic approach developed to study whether remdesivir given during hospitalization for acute COVID will be associated with less Long COVID. The Long COVID outcomes specified here are representative of the NASEM definition, other attempts within the medical literature to define Long COVID,^2,9,12,13,25^ and our developing understanding of the pathobiology of Long COVID as a multisystem disorder.^5,6,14^ These Long COVID are based on ICD-10 diagnoses from medical coding, and other biomarkers as available in the EHR. Additionally, we required that the Long COVID outcomes selected are detectable in our system as differential between persons hospitalized for acute COVID and persons hospitalized for other reasons.

The approach reported here to define Long COVID outcomes has numerous strengths to improve inference in the subsequent research question. First, we pre-specify the outcomes used prior to assessing the exposure in the subsequent experiment, and these decisions are immortalized herein. Second, we address the multiple testing problem associated with a multisystem disorder with over 200 components,^26,27^ with a robust approach which includes specifying a combined primary outcome and individual secondary outcomes for which the family-wise error rate (FWER) is controlled to < 0.05 across the family. Third, we assess the implications for testing the definitions developed in a large dataset in a down sampling experiment simulating a smaller dataset, which predicts which of the secondary outcomes will be robust to the smaller subsequent dataset. In this down-sampling experiment, we show that most of the individual secondary outcomes will be robust in a smaller dataset, with some caveats below. Lastly, we use negative outcomes to improve inference of type 1 errors (false positive). Additionally, we show with negative outcomes and negative exposures that the methods are not likely to show spurious associations. These study design features provide strength of inference for the associations tested in the subsequent research question.

Despite these strengths, several limitations exist. Some features which are clear components of the Long COVID syndrome are not detectable via this approach. For instance, post-exertional malaise (PEM) occurs commonly in Long COVID^13^ and has a pathophysiological basis,^28^ but PEM is not yet captured in the medical terminologies used for structured coding in electronic health records (ICD-10 and SNOMED) to adequately describe this symptom as we previously showed.^29^ Thus, even though PEM is a clear symptom of Long COVID, it could not be included here as an individual secondary outcome and component of the combined primary outcome. Requests should be made for PEM terms to be added to the medical coding ontologies. Other highly prevalent features of Long COVID such as anosmia/dysgeusia^13,30^ may not present commonly to medical attention, so have relatively few events even in such a large dataset. Still other common features of Long COVID such as muscle weakness are common symptoms in the general population, limiting the ability to differentially detect the symptom as a component of Long COVID.^29^ While most of the secondary outcomes will be robust to multiplicity testing in a smaller dataset, some secondary outcomes may be subject to type 2 error (false negative) in the smaller dataset due to low event counts (such as anosmia/dysgeusia) and/or weaker associations (such as anxiety/depression, headache or dysautonomia). These may be due to how common the outcome is in the general population with multiple other causes, which would tend to bias towards the null and weaken the detected association. Thus, the panel of Long COVID outcomes selected here is not comprehensive for defining Long COVID, but is prespecified to answer a specific research question in a subsequent step, i.e. whether remdesivir given during hospitalization for acute COVID is associated with a reduction in subsequent Long COVID. We hope that by pre-specifying these outcomes, while not comprehensive for Long COVID, we can improve inference in this retrospective research.

In stage 2 of this research protocol, we will ask if the use of remdesivir during hospitalization for acute COVID is associated with a reduction in the subsequent development of Long COVID. We will take the cohort of persons hospitalized with acute COVID and assess in this population the exposure of receipt of remdesivir within 3 days of hospitalization versus no receipt of remdesivir to study whether this exposure is associated with less Long COVID by the pre-specified definitions. We will utilize the *a priori* selected primary and secondary Long COVID outcomes and analytic approach specified herein during stage 1. Other details of the analytic approach are equivalent. In addition to analyses performed herein, we will perform the following subsequent additional analyses and sensitivity analyses: negative outcomes testing, negative exposure testing, a different weighting method, sub-group analyses (based on vaccination status, severity and variant era) and sensitivity analyses (considering all RDV exposure including outside of the first 3 days of hospitalization, death as a component of the primary outcome instead of as a competing risk, reinfections instead of primary SARS-CoV-2 infections, and a different window for classification of Long COVID based on 30 days after T_0_). Lastly, exploratory outcomes for hypothesis generation will include the entire set of high-dimensional outcomes.

In summary, the methods presented here are robust for selection of primary and secondary Long COVID outcomes from an EHR dataset, and address challenges inherent in the definition of Long COVID. We show strong associations of the selected outcomes with prior hospitalization with acute COVID, compared to a control group hospitalized for other reasons. These outcomes will be carried forward as *a priori* selected outcomes to a second experiment to test the association of remdesivir with subsequent development of Long COVID.

## Data Availability

All data produced in the present study are available upon reasonable request to the authors.

## Author Contributions

JJH and JDG: Conceptualization; AG, LH, AA, PAM, BJW and JJH: Data curation; AG, LH and AA: Formal analysis; HAA and JDG: Funding acquisition; AG, LH, AA, PAM, MB, EM, BJW, JJH and JDG: Investigation; AG, LH, JJH and JDG: Methodology; HAA: Project administration; BJW, JJH, JDG: Resources; AG, LH, AA and JJH: Software; JJH and JDG: Supervision; AG, LH, PAM, BJW and JDG: Validation; AG and LH: Visualization; AG, LH and JDG Writing – original draft; All authors Writing – review & editing. All authors approve the final submission. JJH and JDG provided supervision and accept responsibility for the integrity and accuracy of the research.

## Data Availability

Data in this report will be made available upon completion of the subsequent research described herein. Data can be obtained at that time by reasonable request to the corresponding author.

