## Supplement for "A protocol for assessment of interventions using a computational phenotype for Long COVID"

Supplemental Information:

Gunjan A., et al. Remdesivir for acute COVID-19 and the association with Long COVID-19: a protocol paper to define Long COVID outcomes.

Supplement 1.pdf Table of Contents:

1. Figure S1: Participant Timeline
2. Figure S2: Analytic Workflow
3. Table S1: Antiviral therapies
4. Table S2: Long COVID Outcomes
5. Table S6: Pre-specified Covariates

Supplement 2.xls Table of Contents:

1. Table S3: High-dimensional Exploratory Outcomes (Diagnosis codes)
2. Table S4: High-dimensional Exploratory Outcomes (Medication codes)
3. Table S5: High-dimensional Exploratory Outcomes (Laboratory thresholds)
4. Table S7: Univariate analysis for top 100 high-dimensional covariates at baseline.
5. Table S8: Covariate Balance
6. Table S9: Primary and Secondary Outcomes
7. Table S10: Exploratory High Dimensional Outcomes in Hospitalized Cohort
8. Table S11: Exploratory High Dimensional Outcomes in Negative Exposure Cohort
9. Table S12: Down-sampling experiment with N=100 replicates.

Supplement 3_protocol.pdf: Protocol version 2.0

- Version date: 04/25/2025
- Sponsor approval date: 04/25/2025
- IRB approval date: 08/01/2025

**Figure S1:** Participant Timeline.

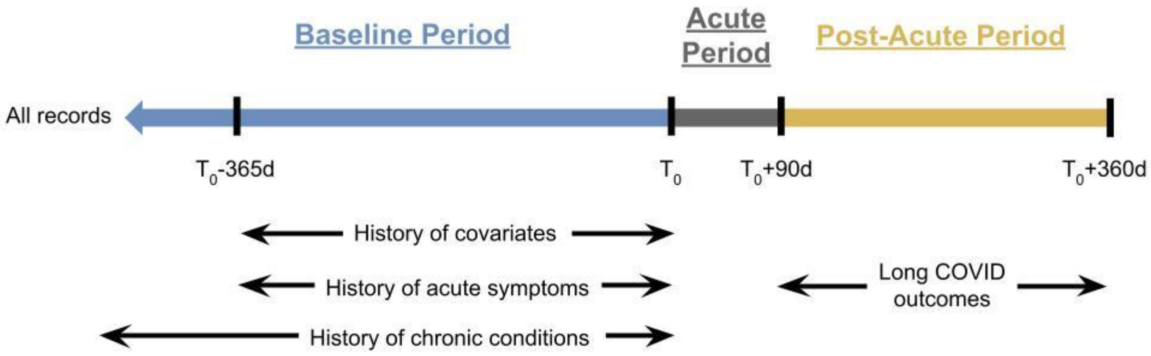


- $T_0$ = admission date (control cohort); earliest of COVID admission or diagnosis (study cohort).
- Outcome-specific cohorts are constructed from individuals without the diagnosis occurring in the relevant baseline period for the diagnosis of interest.
- Outcomes may occur in the acute period, but are not counted until the post-acute period.

**Figure S2:** Analytic Workflow. For each secondary outcome (n=27), a cohort was constructed who did not have the outcome in the relevant baseline period. For the combined primary outcome, the cohort included all persons, but events were counted only if that event did not occur in the relevant baseline period. For the alternative combined outcome, the same cohort was used for the combined primary outcome, though persons who died prior to the outcome assessment period were also included for these deaths to be counted as events.

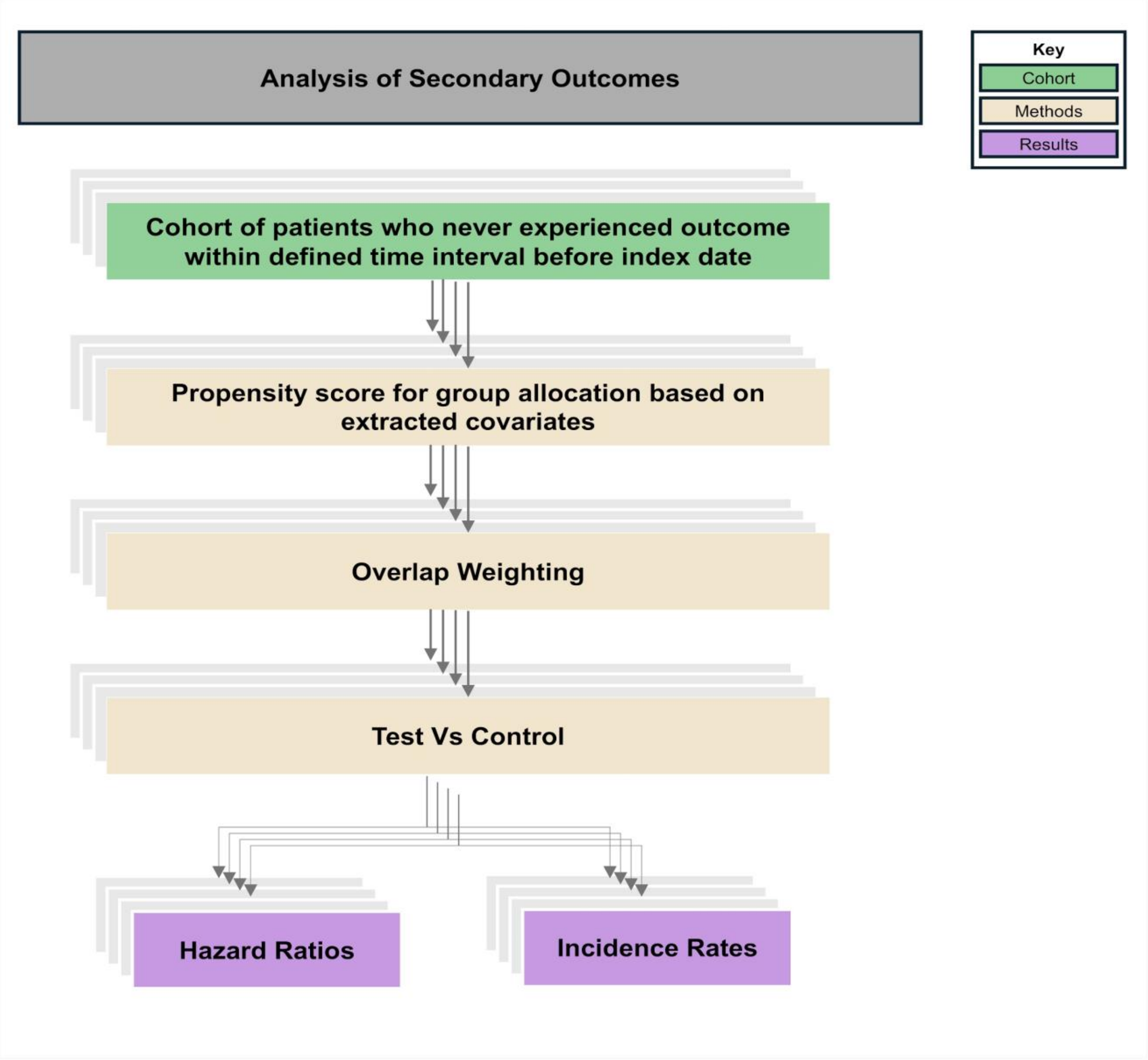

**Table S1:** Antiviral Therapies. Patients were excluded who had received the following COVID-19 antiviral or antibody treatments within 30 days of the index date.

| **Antiviral:** | **RxNorm Codes:** |
|---|---|
| Remdesivir | "2284718" |
| Nirmatrelvir | "2587892","2587899","2599543","2599542","2587898" |
| Molnupiravir | "2603740","2587901","2587906" |
| Monoclonal antibody covid-19 | "2465242","2465249","2592360","2465253","2465248","2463118","2465246", "2571851","2463114","2477902","2479149","2557245", "2592364","2557244","2550731","2550903","2465255","2477854" |

**Table S2:** Long COVID Outcomes. Code and biomarker descriptions for the combined primary Long COVID outcome, individual secondary Long COVID outcomes and negative outcomes. Relevant time frame for baseline period is indicated in the footnote for acute/transient symptoms versus chronic conditions. Biomarker components of the definition and thresholds are given in italics.

| **Organ System** | **Diagnosis** | **ICD-10 codes or Labs & Descriptions:**<br>**(Labs and thresholds given in italics.)** |
|---|---|---|
| **Combined Primary Outcome:** | | |
| **Multisystem** | Long COVID | U09.9 Post-Covid Conditions |
| | Secondary Outcomes | Any incident secondary outcome |
| **Secondary Outcomes:** | | |
| **Cardiac** | Acute coronary syndrome † | I20.0 Unstable angina |
| | | I21x – I23x Acute myocardial infarction [MI] and complications |
| | | *Troponin I or Troponin T > 0.4 ng/mL Biomarker MI* |
| | Arrhythmias † | I44x – I49x Cardiac arrhythmias |
| | | R00.1 Bradycardia, unspecified |
| | Tachycardia | R00.0 Tachycardia, unspecified |
| | Chest pain | R07.2, R07.89, R07.9 Chest pain codes |
| | Heart failure † | I50x Heart failure [HF] |
| | | I11.0, I13.0, I13.2, I27.81 Miscellaneous heart failure |
| | | *Pro BNP > 450 pg/mL or BNP > 100 pg/mL Biomarker HF* |
| | Palpitations | R00.2 Palpitations |
| **Coagulation** | Thromboembolism † | I81, I82x, I80.1x, I80.2x Deep venous thrombosis |
| | | I26x, I27.82 Pulmonary embolism |
| | | I74x, I75x Arterial embolism and thrombosis |
| **Dermatologic** | Hair loss | L65.0, L65.8, L65.9 Hair loss |
| | Skin rash | R21 Rash and other nonspecific skin eruption |
| **Endocrine** | Diabetes mellitus † | E08x – E13x Diabetes mellitus |
| | | O24x Diabetes mellitus in pregnancy, childbirth, and the puerperium |
| | | *HbA1c ≥ 6.5% Biomarker Diabetes mellitus* |

| | | |
|---|---|---|
| | Hyperlipidemia † | E78.0x – E78.5x Hyperlipidemia |
| | | *LDL > 130 mg/dL Biomarker hyperlipidemia* |
| | | *Triglyceride > 150 mg/dL Biomarker hypertriglyceridemia* |
| | Obesity † | E66x Obesity (excluding E66.3) |
| | | O99.21x Obesity complicating pregnancy |
| | | Z68.3x, Z68.4x Body mass index [BMI] ≥30, adult |
| | | *BMI ≥ 30 kg/m2 Biomarker obesity* |
| **Gastrointestinal** | GERD † | K21x Gastroesophageal reflux disease (GERD) |
| | GI Symptoms † | K58.0, K59.1, R19.7 Diarrhea codes |
| | | K58.1, K59.00, K59.01, K59.09 Constipation codes |
| | | R10x (excluding R10.0, R10.2) Abdominal pain |
| | | R11x Nausea and vomiting |
| | | R14.0 Abdominal distension (gaseous) |
| | | R68.81 Early satiety |
| **General** | Fatigue | G93.3x Postviral and related fatigue syndromes |
| | | R53.1 Weakness |
| | | R53.8x Other malaise and fatigue |
| **Kidney** | Kidney dysfunction † | N17x, N19x Acute kidney failure, unspecified kidney failure |
| | | N18.3x – N18.5x Chronic kidney disease, stages 3 – 5 |
| | | N18.6 End stage renal disease |
| | | D63.1, E08.22, E09.22, E10.22, E11.22, E13.22, I12x, I13x, O90.4, Z49x Miscellaneous kidney dysfunction |
| | | *eGFR < 60 cc/min (CKD-EPI 2021) Biomarker kidney dysfunction* |
| **Mental health** | Anxiety / Depression † | F32x, F33x (excluding F32.4, F32.5, F33.4x) Depression |
| | | F40x, F41x Anxiety |
| | | F06.31, F06.32, F06.4, F34.1, F53.0, F94.0 Miscellaneous anxiety and depression |
| | Sleep disorder † | F51x, G25.81, G47x Sleep disorders |

| | | |
|---|---|---|
| **Musculoskeletal** | Joint/Muscle pain | M25.5x, M25.6x Pain or stiffness in joint |
| | | M79.1x, M79.6x Myalgias, muscle pains |
| **Neurologic** | Brain fog | F02x – F03x Dementia |
| | | F04 Amnestic disorder |
| | | R40.0 Somnolence |
| | | F06.7x, F06.8, G31.84, R40.4, R41x Miscellaneous cognitive impairment |
| | Dysautonomia | G90.0x, G90.4, G90.8, G90.9 Disorders of autonomic nervous system |
| | | G90.A Postural orthostatic tachycardia syndrome (POTS) |
| | | G99.0, I95.1, R42 Miscellaneous dysautonomia codes |
| | Headache | G43x – G44x, R51x Migraine and other headache syndromes |
| | Ischemic Stroke/TIA † | G45x Transient cerebral ischemic attack (TIA) |
| | | I63x, I67.82, I69.3x, G43.6x Cerebral infarction, ischemia, and sequelae |
| | Anosmia/dysgeusia | R43x Disturbances of smell and taste |
| **Respiratory** | Cough | R05x Cough |
| | Hypoxemia † | J96.01, J96.11, J96.21, J96.91, R09.02 Hypoxemia codes |
| | Shortness of breath | R06.0x Dyspnea |
| **Negative Outcomes:** | | |
| Neoplasms | Neoplasms † | NEO001-NEO071, NEO073 * Neoplasms benign or malignant |
| Hernias | Hernias † | K40x-K46x Hernias |

† Baseline condition is designated as “chronic” and excluded anytime prior to $T_0$. Otherwise (if not indicated by the † marker), the condition is designated as “acute” and excluded in the 1 year prior to $T_0$.

* Indicated codes are Clinical Classifications Software Refined (CCSR) v.2023.1. Otherwise, all codes are ICD-10.

**Table S3:** Additional criteria for pre-specified Covariates.

| **Feature** | **Feature Descriptions:** *Labs and thresholds given in italics.* |
|---|---|
| History of liver disease/elevated liver function tests * | ICD-10 Diagnosis Codes:<br>K70x – K77x Diseases of liver<br>B15x – B19x Viral hepatitis<br>Laboratory Criteria:<br>*ALT > 5x upper limit of normal or AST > 5x upper limit of normal* |
| Kidney dysfunction * | ICD-10 Diagnosis Codes:<br>N17x, N19x Acute kidney failure, unspecified kidney failure<br>N18.3x – N18.5x Chronic kidney disease, stages 3 – 5<br>N18.6 End stage renal disease<br>D63.1, E08.22, E09.22, E10.22, E11.22, E13.22, I12x, I13x, O90.4, Z49x Miscellaneous codes for kidney dysfunction<br>Laboratory Criteria:<br>*eGFR < 60 cc/min (CKD-EPI 2021) Biomarker CKD* |
| Time since last COVID-19 vaccine *** | In persons with at least 1 vaccine, time since last vaccine was coded as the time in days from the last dose until $T_0$. If no record of vaccine receipt prior to $T_0$, patients were capped (assigned a maximum time) of 1023 days, which is the time from availability of COVID-19 vaccine (12/11/2020) until the last day of follow-up of this study (09/30/2023). |
| Fully vaccinated ** | Centers for Disease Control and Prevention (CDC) CVX Codes:<br>207, 311, 210, 211, 519, 208, 213, 217, 218, 510, 511, 300, 312, 212, 310, 301, 309, 313, 302, 219, 221, 229, 230, 228 |
| Baricitinib/Tocilizumab | RxNorm Medication Codes:<br>'2047232', '612865' |
